# Effect of COVID-19 vaccination on menstrual periods in a prospectively recruited cohort

**DOI:** 10.1101/2022.03.30.22273165

**Authors:** Ee Von Woon, Victoria Male

## Abstract

COVID-19 vaccination protects against the potentially serious consequences of SARS-CoV2 infection, but some people have been hesitant to receive the vaccine because of reports that it could affect menstrual bleeding. To determine whether this occurs, we prospectively recruited a cohort of 79 individuals, each of whom recorded details of at least three consecutive menstrual cycles, during which time they each received at least one dose of COVID-19 vaccine. We find that either dose of the COVID-19 vaccine is associated with a delay to the subsequent period in spontaneously cycling participants (2.3 days after dose 1; 1.3 days after dose 2) but this change rapidly reverses. No change to timing was detected in those on hormonal contraception. We detected no change in menstrual flow associated with either dose of the vaccine, in either spontaneously cycling participants or those on hormonal contraception. We detected no association between menstrual changes and other commonly-reported side effects of vaccination, such as sore arm, fever and fatigue.

## Introduction

As the UK COVID-19 vaccination programme was rolled out to younger participants, the MHRA’s surveillance scheme, Yellow Card, increasingly received reports from people who had noticed a change to their menstrual cycle following vaccination: at 29 March 2022, 39,494 individuals had made such reports to Yellow Card [1]. It is important to note that most people who report such a change following vaccination find that their period returns to normal the following cycle [2] and extensive investigation has found no evidence that COVID-19 vaccination adversely impacts female fertility [3–10]. Nonetheless, people are concerned by these reports. Investigating the potential link between COVID-19 vaccination and menstrual changes is important for maintaining public trust in the vaccination programme and, if a link is found, to allow people to plan for potential changes to their cycles [11].

In response to similar reports to the USA’s vaccine safety surveillance scheme, VAERS, the National Institutes of Health allocated $1.67m (£1.2m; C1.4m) for studies into a possible connection between COVID-19 vaccination and menstrual changes [12] and the first of these studies has recently reported [13]. The authors used prospectively logged data from a menstrual cycle tracking app: 3959 Americans logged at least six consecutive cycles; 2403 of them were vaccinated and 1556 acted as a control group. In individuals receiving only one dose of the vaccine per cycle, the first dose of vaccine had no effect on timing of the subsequent period, while the second dose was associated with a delay of 0.45 days. Individuals who received both doses of the vaccine in a single cycle experienced a 2.32 day delay to their next period. In all groups, cycle lengths returned to normal by two cycles after vaccination.

A Norwegian study used mobile-phone questionnaires to retrospectively solicit reports of menstrual changes from 5688 women aged 18-30 years who had been recruited prospectively to the Norwegian Young Adult Cohort [14]. This study found high levels of variation, even in unvaccinated cycles, with 37.8% of participants reporting at least one difference from normal in their pre-vaccination cycles. However, the study was still able to identify heavier than normal bleeding as a change associated with vaccination, with a relative risk of 1.9 after the first dose of the vaccine and 1.8 after the second dose.

Here, we describe the effects of COVID-19 vaccination on menstrual timing and flow in a cohort of 79 individuals who regularly experience either menstrual periods or withdrawal bleeding as a result of breaks in taking hormonal contraception. These people were recruited before receiving either their first or second dose of the COVID-19 vaccine, and kept a daily record of their vaginal bleeding, and of any vaccine side effects they experienced.

## Methods

### Cohort

253 people who were over 18, have regular periods or withdrawal bleeds and who were planning to receive either their first or their second dose of the COVID-19 vaccine were recruited by advertising on social media and in newsletters with a largely female readership in the UK. Of these, 43 withdrew for reasons including pregnancy, having entered the menopause, having received a diagnosis of a gynaecological condition that they felt was affecting their periods, or not having the time to journal every day (Figure 1). Of the remaining participants, 87 returned their journals and 79 of these journals logged at least three consecutive cycles, including one in which a dose of the vaccine was given. Those who did not return their journals were contacted up to twice to find out why they had not completed their journals: 31 participants responded and 92 were lost to follow up.

**Figure 1.**
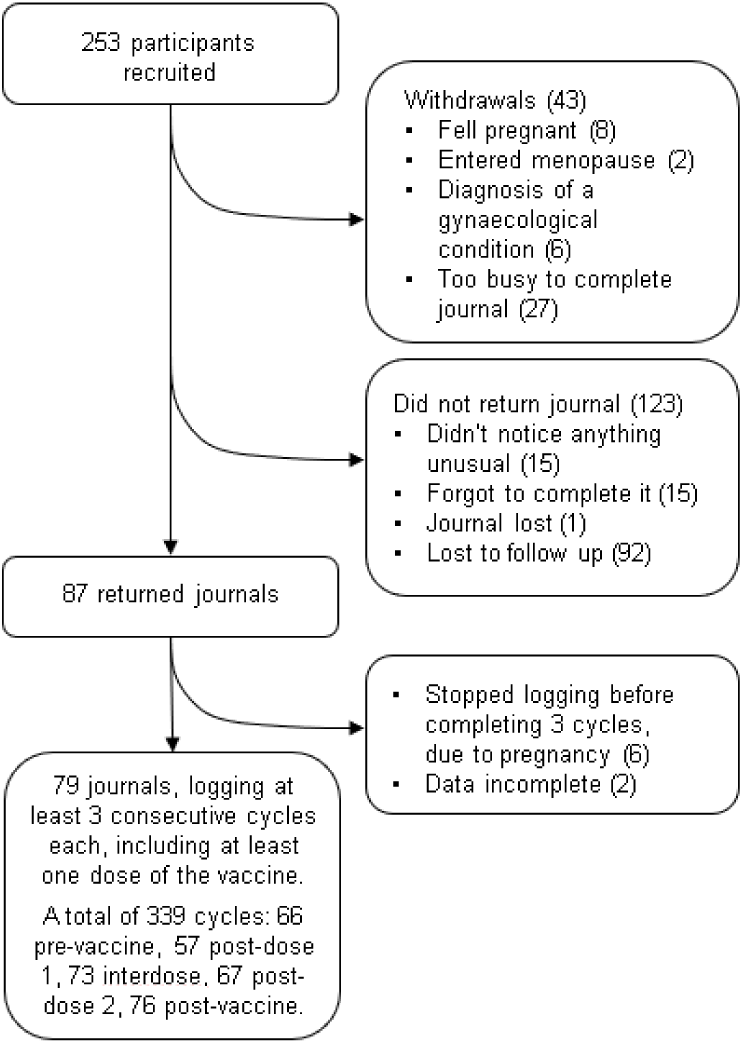
Cohort flowchart.

Journals recorded the participant’s age, reproductive history, use of hormonal contraception during the study period, breastfeeding during the study period and whether they have ever been diagnosed with a menstrual or gynaecological condition (Table 1). Participants then completed a daily journal in which they reported their bleeding as “heavier than usual for this day of my cycle”, “normal”, “lighter than usual for this day of my cycle”, “spotting (normal for me)”, “spotting (not normal for me)” or “no bleeding”. For each cycle, participants also reported whether their period had come on the day they had expected it, and if not how many days early or late it was. Participants noted the day on which they received a dose of the vaccine, which brand they received, whether it was the first or second dose, and for seven days afterwards recorded whether they experienced any of the following: sore arm, fever, fatigue, headache, body aches.

**Table 1.** Participant characteristics.

|  |  |
| --- | --- |
| **Age (Median, IQR)** | 30 (27-35) |
| **Hormonal contraception** |  |
| No hormonal contraception | 63 (79.7%) |
| Combined pill | 7 (8.9%) |
| Progesterone only pill | 3 (3.8%) |
| IUS | 5 (6.3%) |
| Contraceptive patch | 1 (1.3%) |
| **Previous diagnoses** |  |
| Abnormal menstrual bleeding | 4 (5.1%) |
| Heavy menstrual bleeding | 12 (15.2%) |
| Endometriosis | 3 (3.8%) |
| Polycystic ovaries | 7 (8.9%) |
| Uterine fibroids | 5 (6.3%) |
| **Currently breastfeeding** |  |
| Yes | 6 (7.6%) |
| No | 73 (92.4%) |
| **Previous pregnancies** |  |
| 0 | 45 (57.0%) |
| 1 | 15 (19.0%) |
| 2 | 7 (8.9%) |
| 3 or more | 8 (10.1%) |
| Not specified | 4 (5.1%) |
| **Vaccine** |  |
| Pfizer | 65 (82.3%) |
| Moderna | 11 (14.0%) |
| AstraZeneca | 3 (3.8%) |

The study was approved by the Research Governance and Integrity Team at Imperial College London, study number 21IC6988.

### Statistical analysis

The timing of each cycle was designated as “0” when the period or withdrawal bleed started on the expected day, with negative values indicating days early and positive values days late. For each cycle, a “flow score” was calculated by assigning “no bleeding” as 0, “spotting (normal for me)” and “spotting (not normal for me)” as 1, “lighter than usual” as 3, “normal” as 5 and “heavier than usual” as 7, and then totalling the responses for the first seven days of the cycle. Where participants logged more than one cycle in a particular category (for example, pre-vaccine, interdose or post-vaccine cycles) the mean of the timing and flow scores was taken.

For each dose of the vaccine, a “side effect score” was calculated by the total number of days that the participant responded “yes” to each of the side effect questions.

Changes in cycling timing and flow score were assessed using a mixed-effects model, with the Geisser-Greenhouse correction for sphericity, and Tukey’s test for multiple comparisons. Where significant changes were found, sensitivity analyses were carried out by assuming that every participant who had not returned their journal had experienced scores for every cycle with the same distribution as the pre-vaccine cycle scores recorded by those who did return their journals: the distribution was produced by randomly selecting a score from the pre-vaccine cycle data distribution for each missing value.

Associations between side effect, timing and flow scores were assessed using Spearman’s correlation with the Holm-Bonferroni correction for multiple testing.

Analysis was carried out and graphs generated using Graphpad Prism 9.0.0.

## Results

### COVID-19 vaccination is associated with a delay to the next period in spontaneously cycling individuals

Considering only those who were spontaneously cycling, we found that the period after both the first and the second dose of the vaccine came significantly later than expected: compared to the day on which the participant expected their period, pre-vaccine periods occurred on average 0.17 days early, whereas the period following dose one occurred a mean of 2.3 days late (p’ = 0.0045) and the period following dose two occurred a mean of 1.3 days late (p’ = 0.041) (Figure 2a). Periods in interdose cycles and post-vaccination cycles occurred a mean of 0.3 days late and 0.47 days early, respectively, values which were not significantly different from the pre-vaccination average. Therefore, in spontaneously cycling individuals, vaccination was associated with a delay to the subsequent period, but this quickly reversed.

**Figure 2.**
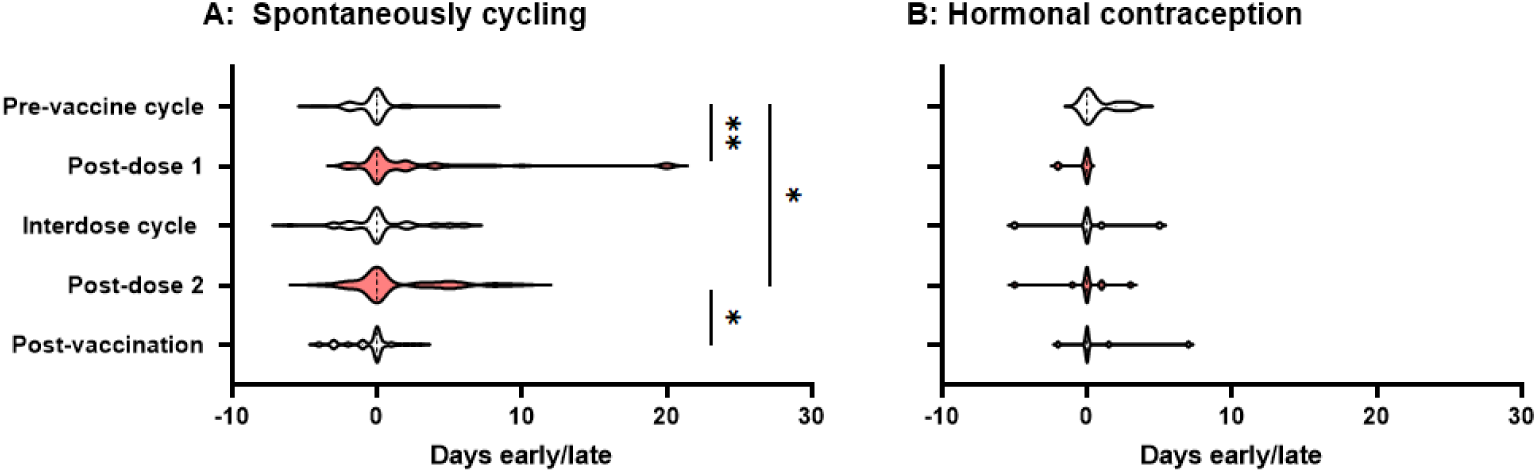
Violin plots showing the distribution by which periods or withdrawal bleeds began in pre-vaccine cycles, the cycle following dose 1 of the COVID-19 vaccine, interdose cycles, the cycle following dose 2 of the COVID-19 vaccine, and subsequent cycles, where “0” denotes the period or withdrawal bleed beginning on the expected day, negative numbers denote days early and positive numbers days late. Data for spontaneously cycling (A) and participants on hormonal contraception (B) are shown. * p’ < 0.05, * * P < 0.01.

Although this finding was in line with that from a large and well-designed study carried out in the USA which also detected delays to the period following a dose of the vaccine [13], the delays we found were somewhat larger. We therefore considered the possibility that those participants who had returned their journals were more likely to have noticed any change than those who did not, inflating the postvaccination changes. To address this possibility, we carried out two sensitivity analyses. We generated distributions consistent with no change occurring and used them to impute the data that we would have expected to see had the participants who responded that they had stopped logging their periods because they had not noticed a change returned their journals: the differences between pre-vaccination cycle timing and post-dose 1 and -dose 2 timing remained significant at p’ = 0.0013 and p’ = 0.011, respectively. Even assuming that none of the participants who failed to return their journals had noticed any change, significant delays to the post-dose 1 period (p’ = 0.0035) and the post-dose 2 period (p’ = 0.045) remained.

Considering only those participants who were taking hormonal contraception, and considering all types of hormonal contraception together due to the small numbers of these participants, we found no effect of vaccination on the timing of the subsequent withdrawal bleed (Figure 2b).

### COVID-19 vaccination is not associated with any change to menstrual flow

We found no significant change to self-reported menstrual flow in the period or withdrawal bleed following vaccination, either in spontaneously cycling participants (Figure 3a), or in those taking hormonal contraception (Figure 3b).

**Figure 3.**
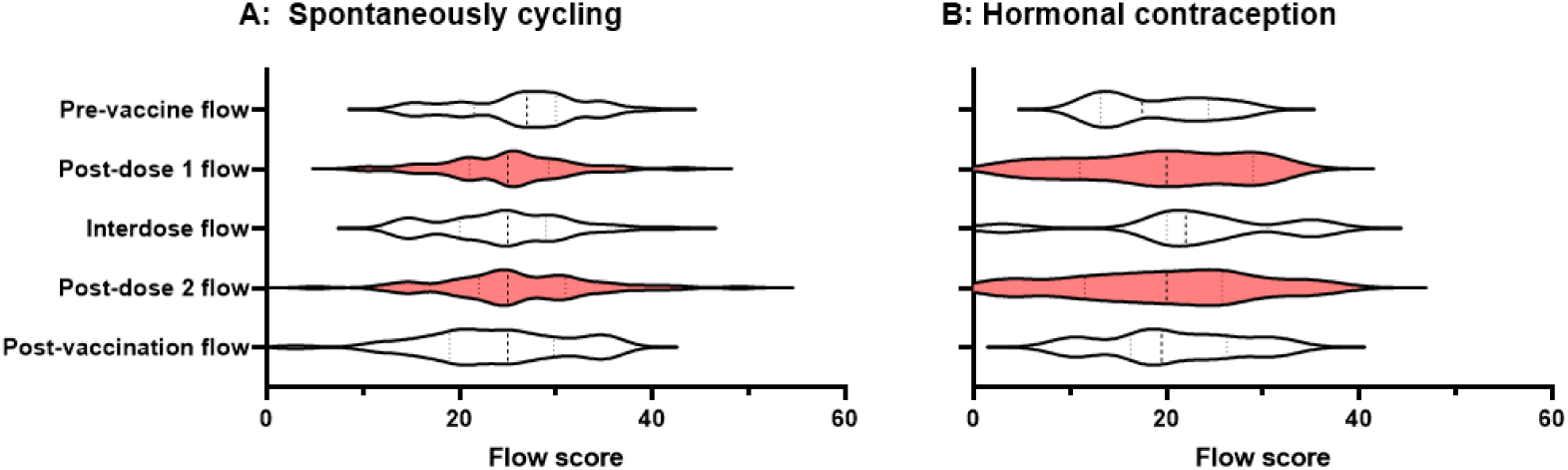
Violin plots showing the distribution of flow scores for periods or withdrawal bleeds in pre-vaccine cycles, the cycle following dose 1 of the COVID-19 vaccine, interdose cycles, the cycle following dose 2 of the COVID-19 vaccine, and subsequent cycles. Data for spontaneously cycling (A) and participants on hormonal contraception (B) are shown.

### Commonly-reported side effects are not associated with menstrual changes

One hypothesis that has been put forward to explain the existence of menstrual changes following vaccination is that immune activation may transiently interfere with the hypothalamic-pituitary-adrenal axis [15,16]. or with immune cells in the lining of the uterus, which control the breakdown and regeneration of this tissue [17]. In either of these cases, we might consider that menstrual changes are not dissimilar to other short-term and reversible side effects of vaccination. Therefore, we might expect menstrual changes to be more likely to occur in those who experienced common side effects.

To explore this hypothesis, we examined correlations between self-reported side effect score and either the timing or the flow score of the subsequent period: no significant correlations were found (Table 2). Therefore, in this small cohort we were unable to find evidence that menstrual changes correlate with other common side effects of vaccination.

**Table 2.** Spearman’s r, p and p’ values are shown for the comparisons indicated.

| Side effect score vs | Dose | Contraception | Spearman's r | p | p' |
| --- | --- | --- | --- | --- | --- |
| Timing | 1 | No | -0.05880 | 0.3575 | 1.00 |
| Flow | 1 | No | 0.1014 | 0.2588 | 1.00 |
| Timing | 2 | No | 0.01375 | 0.4607 | 1.00 |
| Flow | 2 | No | 0.07823 | 0.2870 | 1.00 |
| Timing | 1 | Yes | -0.4648 | 0.3333 | 1.00 |
| Flow | 1 | Yes | -0.4926 | 0.1286 | 1.00 |
| Timing | 2 | Yes | 0.2309 | 0.2268 | 1.00 |
| Flow | 2 | Yes | 0.3159 | 0.1338 | 1.00 |

## Discussion

Here we report that, in a prospectively-recruited cohort, postvaccination periods occurred later than expected after both the first and second dose of the vaccine, and this finding was robust to a sensitivity analysis designed to account for the possibility that participants who noticed a change may be more likely to return their journals. Importantly, periods returned to coming at the expected time in the interdose and post-vaccine cycles. These findings are in line with those from a recent study in the USA, which reported increases in cycle length associated with receiving either the second dose of vaccine, or both doses in the same cycle, but again noted that cycles rapidly returned to normal [13].

In this cohort, we were unable to identify any change in selfreported flow associated with COVID-19 vaccination, and this is in contrast to findings from a Norwegian study, which identified an increase in reports of heavier than normal bleeding after either dose of the vaccine [14]. This cohort was smaller than the Norwegian one, so it is possible that we were not powered to detect a real change in menstrual flow. On the other hand, our participants recorded data in real time, whereas participants in the Norwegian cohort were asked to recall their experiences, so it is also possible that the difference reported in the Norwegian cohort is at least partially impacted by recall bias.

Because our participants were prospectively recruited, this study should be less affected by recruitment bias than survey-based studies [18–20] and because the participants were asked to record their experiences in real-time the data is unlikely to be affected by recall bias. However, there are also a number of important weaknesses. Because the cohort is small, we may not have been able to detect real effects of vaccination on menstrual flow, or real relationships between menstrual and non-menstrual side effects. Some bias is also likely to have been introduced because it is unlikely that the participants who did not return their sheets did so at random, although we did attempt to account for this with a sensitivity analysis. Some bias is also likely to be introduced because “expected date of period” and “menstrual flow” are at least partially subjective, so there is the potential for participant’s expectations of how vaccination will impact their periods to impact their reports.

In conclusion, in this small, prospectively-recruited cohort, we found that both the first and second doses of the COVID-19 vaccine were associated with a small delay to the subsequent period. This delay was not reported in interdose and post-vaccination cycles, suggesting that any changes are temporary. We were unable to detect changes to menstrual flow. These findings are reassuring, but work in larger prospectively-recruited cohorts is needed to more accurately determine how vaccination affects menstrual parameters.

## Data availability

Cleaned, fully anonymised data, together with our analysis files, are available from the Open Science Framework at https://osf.io/upbyg/.

## Supporting information

STROBE checklist for observational studies

## ACKNOWLEDGEMENTS

This study did not receive any specific funding. The investigators were funded by the preterm birth research charity Borne.

## References

1. Medicine and Healthcare Products Regulatory Agency. Coronavirus vaccine—weekly summary of Yellow Card reporting. 25 March 2022. 2021.https://www.gov.uk/government/publications/coronavirus-covid-19-vaccine-adverse-reactions/coronavirus-vaccine-summary-of-yellow-card-reporting

2. Medicines and Health-care Products Regulatory Agency. COVID-19 vaccines: updates for August 2021. https://www.gov.uk/drug-safety-update/covid-19-vaccines-updates-for-august-2021

3. Male V. Are covid-19 vaccines safe in pregnancy? Nat Rev Immunol 2021;21:200–1. doi:10.1038/s41577-021-00525-y

4. Hillson K, Clemens SC, Madhi SA, Voysey M, Pollard AJ, Minassian AM, Oxford COVID Vaccine Trial Group. Fertility rates and birth outcomes after ChAdOx1 nCoV-19 (AZD1222) vaccination. Lancet 2021;398:1683–4. doi:10.1016/S0140-6736(21)02282-0.

5. Morris RS. SARS-CoV-2 spike protein seropositivity from vaccination or infection does not cause sterility. F S Rep 2021; 2:253–255. doi: doi:10.1016/j.xfre.2021.05.010.

6. Orvieto R, Noach-Hirsh M, Segev-Zahav A, Haas J, Nahum R, Aizer A. Does mRNA SARS-CoV-2 vaccine influence patients’ performance during IVF-ET cycle?Reprod Biol Endocrinol 2021;19:69. doi:10.1186/s12958-021-00757-6

7. Bentov Y, Beharier O, Moav-Zafrir A, et al. Ovarian follicular function is not altered by SARS-Cov-2 infection or BNT162b2 mRNA Covid-19 vaccination. medRxiv 2021:2021.04.09.21255195. [Preprint.] doi:10.1101/2021.04.09.21255195

8. Safrai M, Rottenstreich A, Herzberg S, Imbar T, Reubinoff B, Ben-Meir A. Stopping the misinformation: BNT162b2 COVID-19 vaccine has no negative effect on women’s fertility. medRxiv 2021:2021.05.30.21258079 [Preprint.] doi:10.1101/2021.05.30.21258079

9. Aharon C; Lederman M; Ghofranian A et al. In Vitro Fertilization and Early Pregnancy Outcomes After Coronavirus Disease 2019 (COVID-19) Vaccination, Obstetrics Gynecology 2022 doi:

10. Wesselink AK, Hatch EE, Rothman KJ et al. A Prospective Cohort Study of COVID-19 Vaccination, SARS-CoV2 Infection, and Fertility. Am J Epidemiol 2022;, doi:10.1097/AOG.0000000000004713

11. Male V. Menstrual changes after covid-19 vaccination. BMJ 2021;374:n2211 doi: 10.1136/bmj.n2211

12. Eunice Kennedy Shriver National Institute of Child Health and Human Development. Item of interest: NIH funds studies to assess potential effects of COVID-19 vaccination on menstruation. 2021.https://www.nichd.nih.gov/newsroom/news/083021-COVID-19-vaccination-menstruation

13. Edelman A, Boniface ER, Benhar E, et al. Association between menstrual cycle length and coronavirus disease 2019 (covid-19) vaccination: a US cohort. Obstet Gynecol 2022. doi:10.1097/AOG.0000000000004695

14. Tragostad L. Increased occurrence of menstrual disturbances in 18- to 30-year-old women after covid-19 vaccination. [Preprint.] 2022. doi:10.2139/ssrn.3998180

15. Turnbull AV, Rivier CL. Regulation of the hypothalamicpituitary-adrenal axis by cytokines: actions and mechanisms of action. Physiol Rev 1999;79:1–71. doi: 10.1152/phys-rev.1999.79.1.1

16. Karagiannis A, Harsoulis F. Gonadal dysfunction in systemic diseases. Eur J Endocrinol 2005; 152:501–13. doi:10.1530/eje.1.01886pmid:15817904

17. Monin L, Whettlock EM, Male V. Immune responses in the human female reproductive tract. Immunology 2020;160:106–15. doi:10.1111/imm.13136

18. Alvergne A, Kountourides G, Argentieri MA, et al. COVID-19 vaccination and menstrual cycle changes: A United Kingdom (UK) retrospective case-control study. MedRXiv 2021: 2021.11.23.21266709 [Preprint.]

19. Male V. Effect of COVID-19 vaccination on menstrual periods in a retrospectively recruited cohort. MedRXiv 2021: 2021.11.15.21266317 [Preprint] doi:10.1101/2021.11.15.21266317

20. Lee KLM, Junkins EJ, Luo C, Fatima UA, Cox ML, Clancy KBH. Investigating trends in those who experience menstrual bleeding changes after SARS-CoV-2 vaccination. MedRXiv 2021 2021.10.11.21264863 [Preprint] doi:10.1101/2021.10.11.21264863

